# Multimodal Assessment of Peripheral Perfusion for the Early Diagnosis of Sepsis in Critically Ill Patients (MAP-SEPS): A Protocol for an Observational Study

**DOI:** 10.1101/2025.09.28.25336825

**Authors:** Athanasios Chalkias, Ioannis Thivaios, Georgios Karapiperis, Nikolaos Papagiannakis, Fedra Koufaki, Konstantina Katsifa, Athanasios Prekates, Paraskevi Tselioti

## Abstract

**Introduction:** Sepsis-induced organ failure is caused by a dysregulated host response characterized by mitochondrial and microcirculatory abnormalities. Early detection of perfusion deficits is critical to preventing progression to shock and organ failure. While capillary refill time (CRT) and other single-parameter assessments are used, a comprehensive, multimodal evaluation of peripheral perfusion has not yet been applied in clinical settings. The purpose of the MAP-SEPS trial is to ascertain whether such a multimodal approach can enhance early identification of sepsis and organ dysfunction in critically ill ICU patients.

**Methods and analysis:** MAP-SEPS is a prospective observational study enrolling a minimum of 50 adult ICU patients without sepsis on admission. Patients will be monitored over 72 hours using a multimodal protocol that includes clinical (CRT, skin temperature, mottling score, urine output), biochemical (lactate, ScvO□, Pv– aCO□, arterial/interstitial glucose), and near-infrared spectroscopy assessments. Standardized macrohemodynamic monitoring and echocardiography will be performed, along with advanced calculations of venous return dynamics, cardiac efficiency, and arterial/venous resistance. Data will be collected at predefined intervals and analyzed using mixed-effects linear regression models. The primary objective is to assess the predictive value of these hemodynamic and perfusion parameters for early detection of sepsis and organ failure. Secondary outcomes include ICU and hospital length of stay, mechanical ventilation duration, and mortality at 28 and 90 days.

**Ethics and dissemination:** The study has been approved by the Ethics Committee of the General Hospital Tzaneio and complies with the Declaration of Helsinki. Peer-reviewed papers, conference presentations, and clinical seminars will all be used to disseminate the findings, contributing to better bedside evaluation techniques for septic patients.

## Background

Sepsis-related organ dysfunction is caused by a dysregulated host response to infection and involves a variety of interrelated mechanisms, such as cellular injury, metabolic disruptions, coagulation abnormalities, and inflammatory reactions.^1,2^ In recent years, the focus of research on this pathophysiology has shifted from simply identifying the underlying etiology of the disease to examining physiological impairments at all biological levels, including changes in the microcirculation.^3^ The latter are crucial in the development of shock and organ failure and, in the majority of patients—if not all—occur before macrocirculatory alterations during the early phases of sepsis.^4^ The interaction between mitochondrial damage and endothelial cell dysfunction is also heavily influenced by microcirculation.^5–7^ Consequently, microvascular dysfunction may represent a possible window of opportunity for early intervention and slowing of disease progression.

Peripheral perfusion monitoring provides a severity measure that can evaluate hemodynamic incoherence and direct therapy, despite the fact that there are currently no reliable techniques for tracking the patient’s status.^8,9^ This method provides a visual assessment of tissue perfusion by monitoring blood flow in the peripheral microcirculation using physical examination, imaging procedures, and other techniques. Because peripheral perfusion monitoring is easy to access and non-invasive, its use has increased in recent years.

Despite the fact that several tests have been evaluated in clinical practice, such as capillary refill time (CRT),^10,11^ a multimodal assessment of peripheral perfusion using multiple methods or techniques have never been implemented. A multimodal strategy including new approaches, e.g., skeletal muscle microvascular oxygenation (StO_2_), may offer a precise, multifaceted, and real-time evaluation of a patient’s autonomic dysfunction, peripheral vascular reactivity, and tissue perfusion,^12–16^ guiding treatment strategies and permitting a quick therapeutic response.

MAP-SEPS is an observational study that aims to determine whether a multimodal assessment of peripheral perfusion can aid in the early detection of sepsis and organ failure in critically ill patients admitted to the intensive care unit (ICU).

## Methods

### Study design

Critically ill patients admitted to the ICU will be included. The study protocol has been approved by the Ethics Committee of the Tzaneio General Hospital, Piraeus, Greece (16974/22-11-2024). The study will be designed in accordance with the Declaration of Helsinki and will be registered at ClinicalTrials.gov. Written consent will be obtained from patients or their next of kin.

### Study objectives

The primary objective is to investigate the kinetics of various peripheral perfusion monitoring parameters and their prognostic ability for the early detection of sepsis and organ failure in critically ill patients admitted to the ICU. Secondary objectives are to investigate the associations between peripheral perfusion monitoring parameters and hemodynamics, duration of mechanical ventilation, ICU length of stay, in-hospital mortality, 28-day mortality, and 90-day mortality.

### Patient eligibility

Patients fulfilling the following criteria will be included: (a) age ≥18 years; (b) critical illness; and (c) ICU admission. Exclusion criteria include: (a) diagnosis of sepsis at ICU admission; (b) legal incapacity or limited legal capacity; and (c) participation within the exclusion period of another study.

### Circulatory dynamics

All patients will be managed using standard, evidence-based practices according to recent guidelines.^17,18^ A patient-centric approach will be used to provide a quantitative measure of volume state, preload, systemic circuit characteristics, and pump function.

### General hemodynamic assessment

The internal jugular or subclavian vein will be cannulated with a triple-lumen central venous catheter that will be connected to another pressure transducer to measure central venous pressure (CVP) and central venous oxygen saturation (ScvO_2_). The radial artery will be cannulated and connected to a closed-circuit set and transduction system communicating with a monitor, allowing the direct measurement of systolic arterial pressure (SAP), diastolic arterial pressure (DAP) and mean arterial (MAP) pressure. Before making each measurement, we will confirm that transducers are correctly leveled and zeroed,^19,20^ while the system’s dynamic response will be confirmed with fast-flush tests.^21^ Damping will be assessed via repeatedly performed visual inspections of the pressure waveform at the end of a fast flush test. Artifacts will be detected and removed when documented, and when measurements are out-of-range or SAP and DAP are similar or abruptly changed (≥40 mmHg decrease or increase within two minutes before and after measurement). The amount and type of fluids and the dose of vasopressors will be titrated to maintain an individualized mean arterial pressure (MAP) level based on the patient preadmission levels via clinical judgment and dynamic/static tests.

Transthoracic echocardiography will be performed twice daily by a highly experienced echocardiographer who will be blinded to the patient’s identity and study sequence. Left ventricular ejection fraction (LEVF), left ventricular outflow tract (LVOT), LVOT velocity time integral (LVOT-VTI), stroke volume, and cardiac output (CO) will be recorded. All echocardiographic parameters will be calculated from five measurements (regardless of the respiratory cycle) and analyzed retrospectively. Systemic vascular resistance (SVR) will be determined using the CO and MAP [SVR = (MAP – CVP) × 80 /CO].

### Determinants of venous return

The methods of the mean circulatory filling pressure (Pmcf) analogue (Pmca) and related values algorithm have been described in detail before.^5,22–24^ Briefly, based on a Guytonian model of the systemic circulation [CO = *V*R = (Pmcf − CVP) /RVR], an analogue of Pmcf can be derived using the mathematical model Pmca = (a × CVP) + (b × MAP) + (c × CO). In this formula, a and b are dimensionless constants (a + b = 1). Assuming a veno-arterial compliance ratio of 24:1, ‘a’ = 0.96 and ‘b’ = 0.04, reflecting the contribution of venous and arterial compartments, and ‘c’ is a combination of veno-arterial compliance ratio (=0.96) and venous compartment resistance (=SVR × 0.038), with the dimensions of resistance, and is based on a formula including age, height, and weight^25^:

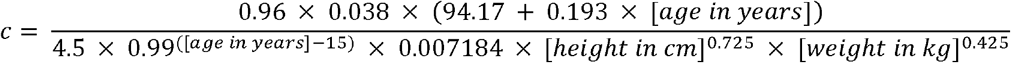

Driving pressure for venous return (VRdP) will be defined as the pressure difference between Pmca and CVP (VRdP = Pmca – CVP). Resistance to venous return (RVR) will be defined as the resistance downstream of Pmca to reflect resistance for venous return and will be calculated as the ratio of the pressure difference between Pmca and CVP and CO [RVR = (Pmca – CVP) /CO]. This formula will be used to describe venous return during transient states of imbalances (Pmca is the average pressure in the circulation and RVR is the resistance encountered to the heart).^26,27^

### Efficiency measures

Efficiency of the heart (E_h_) will be defined as the ratio of the pressure difference between Pmca and CVP and Pmca [E_h_ = (Pmca – CVP) /Pmca] (0 ≤ E_h_ ≤ 1). This equation is proposed for the measurement of heart performance, i.e., how well the heart handles the VRdP in terms of Pmca and CVP.^5,22,24^ A value ~1 reflects a normal heart function with CVP close to 0. During the cardiac stop ejection, right atrial pressure (i.e., CVP) is equal to the Pmca, and E_h_ approaches zero.^5^

Cardiac power [Power = CO × (MAP – CVP) × 0.0022] and power output [CPO = (CO × MAP) /451] will be also calculated. Cardiac power represents the rate of energy input the systemic vasculature receives from the heart at the level of the aortic root to maintain the perfusion of the vital organs in shock states.^28^ Power efficiency (E_power_) will be defined as the ratio between the change in power and the change in Pmca [E_power_ = Δ((MAP-CVP) × CO) × 0.0022 /ΔPmca]. Whereas E_h_ is a static variable, E_power_ dynamically describes the change in cardiac power in relation to the change in power (MAP × CO) and Pmca.^29^

Volume efficiency (E_vol_) will be calculated as the ratio of the pressure difference between Pmca and right atrial pressure (i.e., CVP) and the change in Pmca [E_vol_ = Δ(Pmca-CVP) /ΔPmca] (0 ≤ E_vol_ ≤ 1). Volume efficiency conveys a dynamic variable embodying the efficiency of added fluid, vasopressor, or inotrope in terms of increase in VRdP related to increase in Pmca, and therefore CO and oxygen delivery.^29^

### Other circuit parameters

We will also calculate arterial compliance [C_art_ = SV /(SAP – DAP)], arterial resistance [R_art_ = MAP /(SV × HR)], venous compartment resistance [R_ven_ = SVR × 0.038], and effective arterial elastance (E_a_ = MAP /SV) which is an integrative measure of cardiac afterload that includes steady and pulsatile components.

### Multimodal assessment of peripheral perfusion

The term “multimodal assessment” describes the systematic and comprehensive monitoring of multiple measures to assess particular patient characteristics. Because it is difficult for a single parameter to adequately reflect overall peripheral perfusion in septic patients, this multifaceted assessment has a high clinical reference value.^10^ A comprehensive multi-parameter analysis may correctly assess the association of macrohemodynamics with peripheral perfusion parameters and effectively guide treatment.

The following parameters will be evaluated every four hours for three consecutive days: (a) skin temperature; (b) lactate representing the degree of cell/tissue hypoxia; (c) ScvO_2_ representing systemic oxygen consumption; (d) oxygen extraction ratio (O_2_ER); (e) urine output, (f) venous–arterial carbon dioxide difference (Pv-aCO_2_) representing tissue perfusion and oxygenation imbalance, (g) skin mottling, (h) CRT representing microcirculatory perfusion status, (i) StO□, and (j) arterial blood glucose and interstitial fluid glucose difference (G_A−I_). These perfusion parameters and their kinetics represent many physiological mechanisms of the human body.

## Data collection, monitoring, and management

Clinical data will be obtained through a review of electronic medical records and medical charts. Data analysis will be based on predefined and contemporaneously recorded measurements. The staff will be blinded to measurements until the end of the study and all data are analyzed. An independent Data and Safety Monitoring team will oversee safety, ethical, and scientific aspects of the study. The goal of the clinical data management plan is to provide high-quality data by adopting standardized procedures to minimize the number of errors and missing data, and consequently, to generate an accurate database for analysis. Remote monitoring will be performed to signal early aberrant patterns, issues with consistency, credibility, and other anomalies. Any missing and outlier data values will be individually revised and completed or corrected whenever possible. All researchers will be trained in advance.

The general characteristics of the patients will be recorded, including demographic information, diagnosis, disease severity (e.g., SOFA score — a higher score within 24 h after enrollment indicates more severe organ dysfunction^30^ — and Acute Physiology and Chronic Health Evaluation (APACHE) II score — a higher APACHE II score indicates more severe disease, a poorer prognosis, and a higher mortality rate^31^), and tissue perfusion parameters.

Patients will be subjected to standardized bedside testing at the time of enrollment (H0), H4, H8, H12, H16, H20, H24, H28, H32, H36, H40, H44, H48, H52, H56, H60, H64, H68, and H72 according to predetermined schemes. In terms of perfusion parameters, CRT is the amount of time that passes between a physician applying the proper pressure to a patient’s fingernail bed for 15 seconds in order to precisely remove blood from the nail tip and create a crescent (whitening) underneath the nail bed and the time until the complete restoration of the nail bed’s color.^11^ To reduce measurement error, capillary refill time will be measured three times in a row by two physicians before the mean value is determined.

The skin mottling score is used to conduct a semi-quantitative evaluation of skin mottling at the patient’s bedside and represents the size of mottled regions on the patients’ knees and thighs when they lie supine with their legs stretched out and flush with their hearts.^32^ This system uses a scoring system with a range of 0 to 5; a score of 0 indicates that there is no mottling, while scores 1 through 5 indicate progressively more severe mottling. In particular, a score of 1 means that the mottling is limited to the knee’s center, whereas a score of 2 means that it extends to the kneecap’s perimeter. A score of four means that the mottling has expanded to the ends of the thigh and calf, whereas a score of three means that it is above the knee but does not extend past the mid-thigh and calf. Lastly, mottling that has extended to the ankle and groin is represented by a score of 5.

Using a central venous catheter, a central venous blood sample will be drawn from the patient, and a blood gas analyzer will measure the amount of ScvO_2_ in the blood sample. Additionally, Pv-aCO_2_ will represent the carbon dioxide difference, which is determined by subtracting the partial pressure of carbon dioxide in the arterial blood from that in the central venous blood. This information will be derived from the simultaneous collection of arterial and central venous samples.

Using a near-infrared spectroscopy (NIRS) instrument, the skeletal muscle microvascular oxygenation patterns of the right vastus lateralis will be assessed. After the area has been carefully shaved and dried, the probe will be affixed to the skin above the vastus lateralis muscle (about 12 cm above the kneecap) using adhesive tape and an elastic bandage. Oxygenated and deoxygenated hemoglobin/myoglobin concentrations (oxy-[Hb/Mb] and deoxy-[Hb/Mb], respectively) can be measured using NIRS.^33^ Additionally, tissue oxygen saturation (StO□ = oxy-[Hb/Mb]/total - [Hb/Mb]) and total heme concentration (total-[Hb/Mb] = oxy-[Hb/Mb] + deoxy-[Hb/Mb]) will be computed.^33^ The deoxy-[Hb/Mb] signal, which was measured from resting baseline to fatigue, has been regarded as a surrogate marker of fractional oxygen extraction in the microcirculation, representing the balance between oxygen delivery (DO_2_) and consumption (VO_2_).^34^

The finger-pricking technique and a standard glucose meter will be used to collect interstitial fluid glucose at the same time as arterial blood gas analysis and arterial blood glucose measurement.^35^ The interstitial fluid glucose difference will be calculated as the difference between the two measurements.

Sepsis will be diagnosed via clinical assessments and basic laboratory tests, cultures, imaging studies, and sepsis biomarkers, as indicated, in conjunction with SIRS criteria [tachycardia (heart rate >90 beats/min), tachypnea (respiratory rate >20 breaths/min), fever or hypothermia (temperature >38 or <36 °C), and leukocytosis, leukopenia, or bandemia (white blood cells >1,200/mm^3^, <4,000/mm^3^ or bandemia ≥10%)] (https://www.mdcalc.com/calc/1096/sirs-sepsis-septic-shock-criteria).^17,18^

Sepsis-related, non-compensatory tachyarrhythmia will be managed with intravenous landiolol, titrated based on systemic hemodynamics and tissue perfusion.^6,36,37^ Macrohemodynamic evaluation will also include analysis of the systolic–dicrotic pressure difference, which may indicate a reduced rate of pressure change over time (dP/dt_max) and reflect the degree of coupling between myocardial contractility and afterload.^38^ Tissue perfusion will be assessed using the previously described multi-parameter analysis of peripheral perfusion.

## Sample size and predefined statistical analysis plan

No previous studies in the literature have been found that assess peripheral perfusion with a multimodal approach to support sample size calculation. We chose to include a minimum of 50 individuals because we expected that this number could reveal important associations and generate results to be used for sample size estimation in future large scale studies.

Data will be presented as mean (SD), median (interquartile range) or frequency and proportion. Baseline characteristics of the patients will be reported using the Wilcoxon rank-sum test for continuous variables and the χ^2^ test or Fisher’s exact test for categorical variables, when appropriate. Mixed effects linear regression will be used to assess the longitudinal differences.

The level of statistical significance will be set at 0.05, and all tests will be two-tailed. The Benjamini-Hochberg false discovery rate correction will be used to account for the multiple comparisons necessitated by the study protocol and the exploratory nature of this analysis. Analyses will be performed using R software (version 4.3.0, The R Foundation for Statistical Computing Platform, 2022).

## Discussion

Effective sepsis management requires early detection and prompt response, as delays greatly increase morbidity and mortality among severely ill patients. Traditional systemic hemodynamic parameters may fail to reflect early tissue hypoperfusion, necessitating a multimodal assessment of peripheral perfusion that integrates systemic, regional, and microcirculatory markers to improve diagnostic accuracy and guide therapy.^39^

Clinical examination remains a valuable tool in sepsis. Peripheral skin temperature gradients, especially between the core and extremities, reflect sympathetic vasoconstriction. A central-to-toe temperature difference of >4°C has been associated with poor tissue perfusion and adverse outcomes.^40^ Capillary refill time longer than 3 seconds is associated with elevated lactate levels and increased 28-day mortality in septic shock.^41^ Mottling, assessed via a semi-quantitative score, has similarly been shown to predict mortality. For example, Ait-Oufella et al. found that a mottling score ≥3 at 6 hours was associated with over 60% mortality versus <10% for a score of 0.^32^ Urine output, though nonspecific, reflects renal perfusion and sustained oliguria (<0.5 mL/kg/h) is independently associated with ICU mortality.^42^

Serum lactate remains a cornerstone biomarker in sepsis and reflects tissue hypoperfusion. Elevated lactate (>2 mmol/L) is associated with increased mortality, and failure to clear lactate by at least 10% within 6 hours represents a poor prognostic sign.^43^ However, lactate levels may also be elevated due to adrenergic stimulation or impaired clearance, necessitating complementary assessment.

Central venous oxygen saturation reflects global oxygen balance. Values <70% suggest inadequate oxygen delivery, whereas paradoxically high values (>80%) with rising lactate may indicate impaired tissue oxygen extraction or mitochondrial dysfunction.^44^ The oxygen extraction ratio provides further insight into the tissue’s ability to utilize oxygen. Elevated O□ER (>30%) in the context of hypotension or organ dysfunction indicates increased tissue demand or insufficient perfusion.^45^

The venous-to-arterial carbon dioxide difference (Pv-aCO□) is a sensitive marker of low-flow states and inadequate perfusion. A Pv-aCO□ greater than >6 mmHg correlates with impaired cardiac output and worse outcomes even when ScvO□ is normal.^46^ After initial resuscitation, patients with Pv-aCO□ >6 mmHg have nearly double the 28-day mortality compared to those with lower ratings.^47^ Furthermore, the ratio of Pv-aCO□ to arteriovenous O□ content difference (Pv-aCO□/C(a-v)O□) >1.4 has been proposed as a surrogate for anaerobic metabolism.^48^

Near-infrared spectroscopy allows for the non-invasive assessment of StO□. Studies have demonstrated significantly lower StO□ levels in septic patients compared to controls, often preceding changes in systemic parameters.^49^ Creteur et al. found that patients with baseline StO□ <75% experienced higher mortality, and impaired StO□ recovery after vascular occlusion testing was associated with worse outcomes.^50^ A meta-analysis confirmed that decreased StO□ and a delayed reperfusion slope predict ICU mortality in sepsis.^51^

Impaired microvascular perfusion disrupts cellular glucose delivery. The arterial-to-interstitial glucose gradient, measurable via microdialysis or continuous glucose monitoring, increases in sepsis and reflects regional hypoperfusion and cellular metabolic stress.^35,52^ Indeed, tissue glucose levels may drop to <2 mmol L^-1^ in septic shock despite normal systemic glucose, indicating impaired substrate delivery and utilization.^53,54^ Combining microcirculatory and metabolic indicators such as NIRS and glucose gradients may enable earlier detection of tissue hypoxia and better individualization of therapy.

## Ethics and dissemination

The study was approved by the Ethics Committee of the Tzaneio General Hospital (16974/22-11-2024) and adheres to the Declaration of Helsinki, the International Conference on Harmonization guideline for Good Clinical Practice, and applicable local regulatory bodies. Written informed consent will be obtained from the patient or the patient’s legally authorized representative prior to the initiation of any study procedure. The final study dataset will be available to investigators. Datasets will be stored for 20 years following trial completion. Study results will be submitted to peer-reviewed journals, and the results will be presented at one or more scientific conferences, with an expected timeframe for publication between 2026 and 2027.

## Data availability

Data will be made available upon request after publication through a collaborative process. Researchers should provide a methodically sound proposal with specific objectives in an approval proposal. Please contact the corresponding author for more information.

## Acknowledgements

Nothing to acknowledge.

## Authors contributions

AC contributed to study design, protocol draft, review and editing of the manuscript. IT, GK, FD, KK, AP, and PT contributed to protocol draft and manuscript editing. NP contributed to protocol draft, biostatistical planning and editing of the manuscript. All authors revised the manuscript for important intellectual context and gave final approval for publication.

